# Medtronic Percept^™^ Recorded LFP Pre-Processing to Remove Noise and Cardiac Signals From Neural Recordings

**DOI:** 10.1101/2025.03.17.25324121

**Authors:** Zachary Sanger, Steffen Ventz, Robert McGovern, Tay Netoff

## Abstract

Chronic brain sensing devices, such as the Medtronic Percept™ or Neuropace RNS system, record local field potentials (LFPs) that may be vulnerable to noise from hardware limitations, environmental factors, movement, stimulation, cardiac signals, and analytical procedures. Although onboard hardware filters can attenuate some noise, additional processing is often required. Here we demonstrate that cardiac artifacts significantly alter the power spectral density (PSD) of neural activity within the theta (4– 8 Hz), alpha (8–12 Hz), and beta (12–30 Hz) bands. We introduce a time-domain template subtraction method specifically designed to remove QRS complex cardiac artifacts. Separately, we describe techniques for transforming time domain data to the frequency domain and mitigating transient artifacts by estimating background neural activity—either through window rejection based on PSD characteristics or via principal component analysis. Finally, we present an approach to isolate oscillatory neural activity by subtracting the aperiodic 1/f component from the power spectrum by fitting the FOOOF logarithmic function. While filter selection must be tailored to the specific device and participant environment to avoid over-filtering, these noise mitigation strategies are crucial for ensuring the integrity of LFP recordings.

## Introduction

### Local Field Potentials and Sources of Noise

Historically, invasive neuromodulation has employed open-loop therapy, wherein patient feedback during surgery and subsequent neurologist visits informs clinicians in setting static stimulation parameters. Recently, neuromodulation therapies such as Neuropace’s RNS and Medtronic’s adaptive Deep Brain Stimulation (aDBS) systems have leveraged local field potentials (LFPs) recorded near implanted leads to capture changes in neural activity related to patient state or stimulation response. Neural oscillations within broadband LFP signals, e.g., the beta band in Parkinson’s disease [1–4], may serve as valuable quantitative biomarkers that can be exploited in closed-loop therapeutic approaches. However, closed-loop neuromodulation can be susceptible to unwanted noise signals that may vary over time.

While LFPs contain neural signals, they are susceptible to noise from both the recording hardware and the environment. LFP recordings are measured as the voltage difference between two electrode contacts, often on the same implanted lead to isolate local neural activity. For noise sources distant from the electrodes, the signals are nearly identical at both contacts; subtracting them thus removes this common noise, a process known as common mode rejection. This technique effectively reduces cardiac artifacts and powerline interference generated outside the brain.

Implantable recording hardware generally mitigates signal drift by filtering out low-frequency components—referred to as high-pass filtering by engineers—and attenuating signals such as 60 Hz power line noise (or 50 Hz in Europe) using notch filters. When digitizing a signal, the highest frequency that can be accurately represented is half the sampling frequency, known as the Nyquist frequency. Frequencies above the Nyquist limit are folded back into the lower frequency range, a phenomenon termed aliasing, which corrupts the signal. Consequently, anti-aliasing low-pass filters are applied to eliminate frequencies near or above the Nyquist threshold prior to digitization.

When converting these analog differential signals—already filtered by hardware—into a digital format, both sampling and digitization noise can affect the fidelity of the local field potential recordings. Sampling noise arises from measuring voltage at discrete time intervals, potentially impacting signals with frequencies near the Nyquist limit. To accurately reconstruct a signal, sampling at a rate approximately ten times the frequency of interest is advisable; however, higher sampling rates demand increased energy and storage, posing practical limitations.

Digitization, or quantization, noise is introduced by the rounding errors inherent in converting the analog signal to digital numbers. Factors such as the analog-to-digital converter’s (ADC) resolution and its slew rate, which defines how quickly the electronics respond to changes in input voltage, determine the accuracy and precision of this conversion. Notably, digitization noise is uniformly distributed across the frequency spectrum.

Once digitized, the signal can be processed using a variety of data analysis techniques to mitigate different noise sources. Transient artifacts, such as movement artifacts, are often addressed at the software level. Nevertheless, many LFP recordings require additional filtering to remove residual powerline noise, cardiac and stimulation artifacts, background activity (1/*f* noise), digitization noise, and signal changes due to variations at the electrode interface over time. Software processing methods can be employed to isolate specific LFP components that, although not strictly noise, might otherwise confound the analysis of neural signals

### Addressing Noise in Local Field Potentials

We have included LFPs recorded from the anterior nucleus of the thalamus (ANT) of refractory epilepsy patients to illustrate different types of noise that can be observed within LFP recordings. These data also demonstrate that human data recorded from commercially available FDA approved systems may still require noise rejection.

Noise does not necessarily corrupt recorded LFPs; rather, different recording setups exhibit varying susceptibilities to noise sources. Properly addressing artifacts and noise ensures that the recorded data accurately represents neural activity. In this paper, we outline considerations and approaches for mitigating artifacts and extraneous neural signals in LFP recordings. These methodologies may benefit clinicians and scientists analyzing LFPs recorded from implanted devices such as Medtronic’s Percept™.

In this paper, we describe a method for eliminating cardiac artifacts that may persist in certain patient recordings, approaches for transforming LFPs from the time domain to the frequency domain, strategies to mitigate movement artifacts or large transient changes in the data (which may also be neural in origin), and an approach for removing or characterizing the background noise, commonly referred to as 1/*f* noise.

## Methods

### Recording with Medtronic’s Percept™

The Medtronic Percept™ PC and RC systems offer commercially available features that support various recording paradigms in both clinical and home settings, with or without stimulation. Using the clinician programming tablet, Percept™ DBS systems can capture LFPs via BrainSense™ Survey, BrainSense™ Survey Indefinite Streaming, and BrainSense™ Streaming modes. These recording modes, designed for clinical use, sample data at 250 Hz. In contrast, at home, an LFP power measurement is saved every 10 minutes within a predefined 5 Hz band. BrainSense™ Survey can be used in the clinic to record roughly 20 seconds of multiple bipolar signals on the same lead with stimulation off. Indefinite Streaming limits this survey to the 3 ring pairs on each lead, but allows for an extended recording, primarily limited by file size. Here, the data shown was collected using BrainSense Streaming to allow for recording during stimulation, but this only allows for one bipolar recording pair per lead.

In our trial, LFPs are segmented into 1-minute recordings capturing responses during both stimulation on and off states. All recordings are acquired using the BrainSense Streaming mode, with the baseline (stimulation off) condition achieved by setting the stimulation amplitude to 0 mA.

### Subjects

Data in this paper is presented from 11 participants in an ANT Deep Brain Stimulation (DBS) optimization clinical trial for treatment of refractory epilepsy. All recordings were done in accordance with an Institutional Review Board (IRB)-approved clinical study (#NCT05493722) at the University of Minnesota. Participants received a Medtronic Percept™ system with either Legacy (3387/3389) or Sensight™ (B33005/B33015) leads implanted in the ANT. The study investigates the ANT’s response to various stimulation parameters by testing multiple settings around the clinical parameters. For the trial analysis, we assume a stationary neural response during each 1-minute segment of stimulation-off (baseline) and stimulation-on testing for each parameter setting. Here, we present example recordings to illustrate these artifacts and signals, along with long-term data on these artifacts observed in participants’ LFP recordings over time.

## Results

In this work, we demonstrate how artifacts were removed from LFP signals recorded with Medtronic’s Percept™ device. Specifically, we address the removal of cardiac artifacts—which can distort estimates of spectral power in the lower frequency bands, transforming LFPs into the frequency domain, removal of transient signals that intermittently affect the spectral broadband power, and removal of the background aperiodic signal, often referred to as 1/*f* background noise.

### Time Domain Removal of ECG QRS Artifacts

In all participants with Medtronic Legacy (3387/3389) leads, cardiac artifacts were observed in both ANT LFP recordings at the first neurology follow-up visit. We have observed cardiac artifacts and/or transient signals in 7 of 11 participants enrolled in our study. Among eight participants with Sensight™ (B33005/B33015) leads, only 50% exhibited an observable QRS complex in the ANT LFP recordings. Notably, in three of the four Sensight lead participants with ECG artifacts, these were not detected until approximately the 15th month of follow-up. Further details regarding the implanted pulse generator (IPG) subclavicular placement, lead type, and the timing of initial ECG artifact observation in ANT LFP recordings are provided in Table 1.

**Table 1:** Bilateral Anterior Nucleus of Thalamus Participant Population Implant Details.

| ID: | Device | Placement | Additional Devices | Lead Type | ANT ECG Artifact | Days Since Implant (QRS Visually Observed) |
| --- | --- | --- | --- | --- | --- | --- |
| 1 | Replacement to Percept™ PC | Right |  | Legacy | Both | 1 (Visit 1) |
| 2 | Replacement to Percept™ PC | Left | RNS | Legacy | Both | 1 (Visit 1) |
| 3 | Percept™ PC | Right |  | Legacy | Both | 50 (Visit 1) |
| 4 | Percept™ PC | Left |  | Sensight™ | Both | 1 (Visit 1) |
| 5 | Percept™ PC | Right |  | Sensight™ | Both | 498 (Visit 4) |
| 6 | Percept™ PC | Right |  | Sensight™ | Right | 420 (Visit 3) |
| 7 | Percept™ PC | Right |  | Sensight™ | Right | 493 (Visit 7) |
| 8 | Percept™ PC | Right |  | Sensight™ | None (864) |  |
| 9 | Percept™ PC | Right | VNS - off | Sensight™ | None (753) |  |
| 10 | Percept™ PC | Right | VNS - on | Sensight™ | None (647) |  |
| 11 | Percept™ PC | Right |  | Sensight™ | None (260) |  |

Some participants exhibit varying intensities of the ECG QRS wave signal, as illustrated in Figure 1A. In this participant, the QRS complex amplitude is considerably smaller in the left ANT recording compared to the right. To address this artifact, we removed the QRS complex using a template subtraction method [5–7]. For template matching of the QRS complex, we first band-pass the LFP time-domain signal between 5 and 30 Hz to isolate the cardiac signal from low and high frequency neural and noise components. Next, we identify the R wave event times using MATLAB’s `findpeaks` function, setting the “MinPeakDistance” to 0.5×250 samples/second and the “MinPeakProminence” to 2.5 times the standard deviation of the filtered signal. Since the R wave may be inverted depending on electrode location, it may be necessary to multiply the signal by -1 to detect the positive R wave peak.

**Figure 1:**
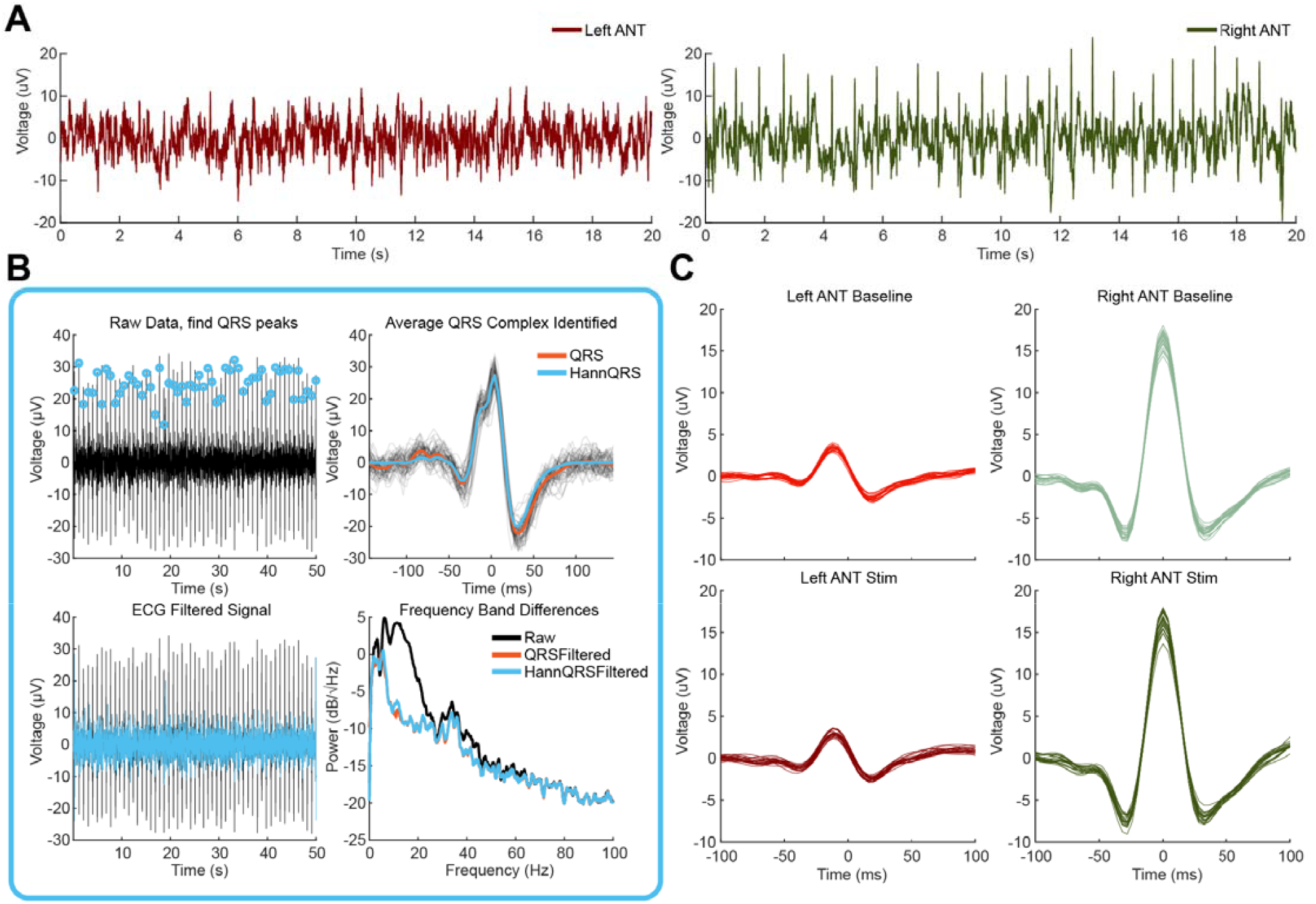
Screening Neural Recordings and Removing Cardiac Artifact. **A) Participant 6 raw Percept™ recorded ANT LFPs from the left and right hemisphere. Note the right hemisphere shows a greater QRS than the left and the left may be hard to even perceive it exists. B) ECG template subtraction workflow. The raw LFP signal in the hemisphere with greater QRS artifact is bandpassed between 5-30 Hz and used to identify peaks. An average QRS complex is determined, windowed using a Hanning window, and subtracted at the locations of the QRS in the original data. The ECG filtered data PSD shows the dom nance of ECG artifacts in the theta, alpha, and beta activity. C) Participant 6 Differences in ECG artifact between hemispheres. The left hemisphere in baseline and stimulation recordings had a much smaller QRS than the right hemisphere. This small QRS can still be extracted by utilizing the QRS locations identified in the right hemisphere.**

Once the peaks are identified, a QRS complex template is constructed by averaging the QRS windows surrounding the detected peaks, as illustrated in Figure 1B. To capture the Q and S waves, a minimum window size of 18 samples prior to and 36 samples following the R wave is required. To avoid introducing abrupt changes during template subtraction, the QRS template is multiplied by an equal length Hanning window. This approach forces the edges of the template to zero while preserving the central components, thereby preventing the introduction of large clicks at the template boundaries. To ensure the R wave is not attenuated by an asymmetrical windowing function, a symmetrical Hanning window is employed, with a minimum of 36 samples before and after the R wave.

As shown in Figure 1B, the ECG QRS signal significantly contributes to power in the theta, alpha, and beta band frequencies of the power spectral density (PSD), which are markedly reduced following artifact removal. This is why removing cardiac artifacts prior to transforming the data into the frequency domain is preferable. PSD plots represent squaring the magnitude estimated at different frequencies when transforming LFPs into the frequency domain. Often, the power versus frequency data in a PSD is converted into deciBels (dB) for better visualization and so the data resembles a normal distribution due to the 1/*f* background noise.

Lastly, since the cardiac signals contaminating both hemispheres are time-locked, the R wave event times estimated from the hemisphere with the larger cardiac signal can be used to remove QRS complexes from both hemispheres. In Figure 1C, the QRS complexes from both baseline and stimulation conditions (from the same participant shown in Figure 1A) were extracted from one minute of baseline and one minute of stimulation data. The R wave event locations from the right hemisphere were used to extract QRS complexes from both hemispheres. This example demonstrates that even subtle ECG artifacts in one hemisphere may be present and can be effectively removed using R wave detection from the hemisphere with the more prominent artifact.

In some participants, we observed variations in QRS complex amplitude between stimulation and baseline recordings. To account for these differences, separate QRS templates are generated for each recording state within BrainSense Streaming.

### Visualizing Local Field Potentials in the Frequency Domain

Specific frequency band activity, such as the beta band (12-30Hz) observed in Parkinson’s disease patients [1–4], can be useful data for assessing the patient state or efficacy of therapy. To visualize different neural oscillation frequency bands, such as theta (4-8Hz), alpha (8-12Hz), beta (12-30Hz), slow gamma (30-50Hz), or high gamma (50-200Hz), the time domain LFPs must be transformed to the frequency domain. In neural signal analysis, the Fourier and Wavelet transforms are popular frequency transforms depending on the application. Here, we share important considerations when using the Fourier transform to extract frequency band activity from time domain LFPs.

### Frequency Transform

LFPs can be transformed into the frequency domain using the entire recording or by transforming smaller overlapping segments. These segments can be averaged together to create a PSD of the entire recording or displayed individually to show how the PSDs change over time, called a spectrogram. The spectrogram can be useful for observing changes in frequency content over time, or to isolate windows of data that are different. The Fourier transform assumes stationary data, meaning the data is consistent and unchanging throughout the segment being transformed, which is often not true in neural recordings.

Transforming neural signals from the time domain to the frequency domain is performed by the Discrete Fourier transform (DFT). The DFT decomposes the LFP by comparing it to sine and cosine waves, which can then be used to calculate the amplitude, frequency, and phase of oscillations in the LFP. If particular sine and cosine frequencies account for the majority of the LFP signal, these frequencies appear as peaks on a PSD plot. The length of the DFT (`nfft`) and sampling rate (*fs*) determines all the sine and cosine frequencies which will be used to transform the LFP signal.

The DFT treats the data at the beginning and end of the segment as temporal neighbors, as if on a loop. If the data at the ends do not match, it causes a “click” in the data, which can contribute a significant spectral component that is confounded with the spectrum of the neural data, called spectral leakage. Spectral leakage can be mitigated by applying a windowing function to the LFP segment to prevent these clicks at the ends of the data segment. Instead, the windowed segmented data appears as though no signal exists at the edges and data is only preserved in the middle of the LFP segment. To better retain the information at the edges of the segments, segments can be stepped by half the window length and averaged. An alternative method for retaining information is the multitaper transform, which uses multiple windows in that same segment to include information throughout the whole segment and then these windows are averaged together [8,9].

### Hanning Windowing

The Hanning window is often the default windowing function, which forces the data segment edges to zero. The length of the Hanning window is set to the data segment length. Data segment length, or window length, is chosen based on its PSD frequency resolution impact broken down into: 1) the lowest frequency observed based on the length of the segment of data, 2) the minimum frequency resolution that can be observed based on the length (`nfft`) of the DFT, often calculated using the Fast Fourier Transform (FFT) for computational efficiency, and 3) the frequency resolution due to smoothing of the PSD by the windowing function.

Software implementation of the DFT requires an LFP recording and selection of the window length, Fourier transform length (`nfft`), and the segment overlap. Often, the `nfft` sample lengths will be an integer of base 2 to take advantage of the computational efficiency of the Fast Fourier Transform. If the window length is shorter than the Fourier transform length, the windowed data will be zero padded symmetrically on both ends of the segment to match the length of the `nfft`. Because the input data is much longer than the window length, the overlap defines how to divide up this data into multiple windows, as shown in the middle graph of Figure 2D.

**Figure 2:**
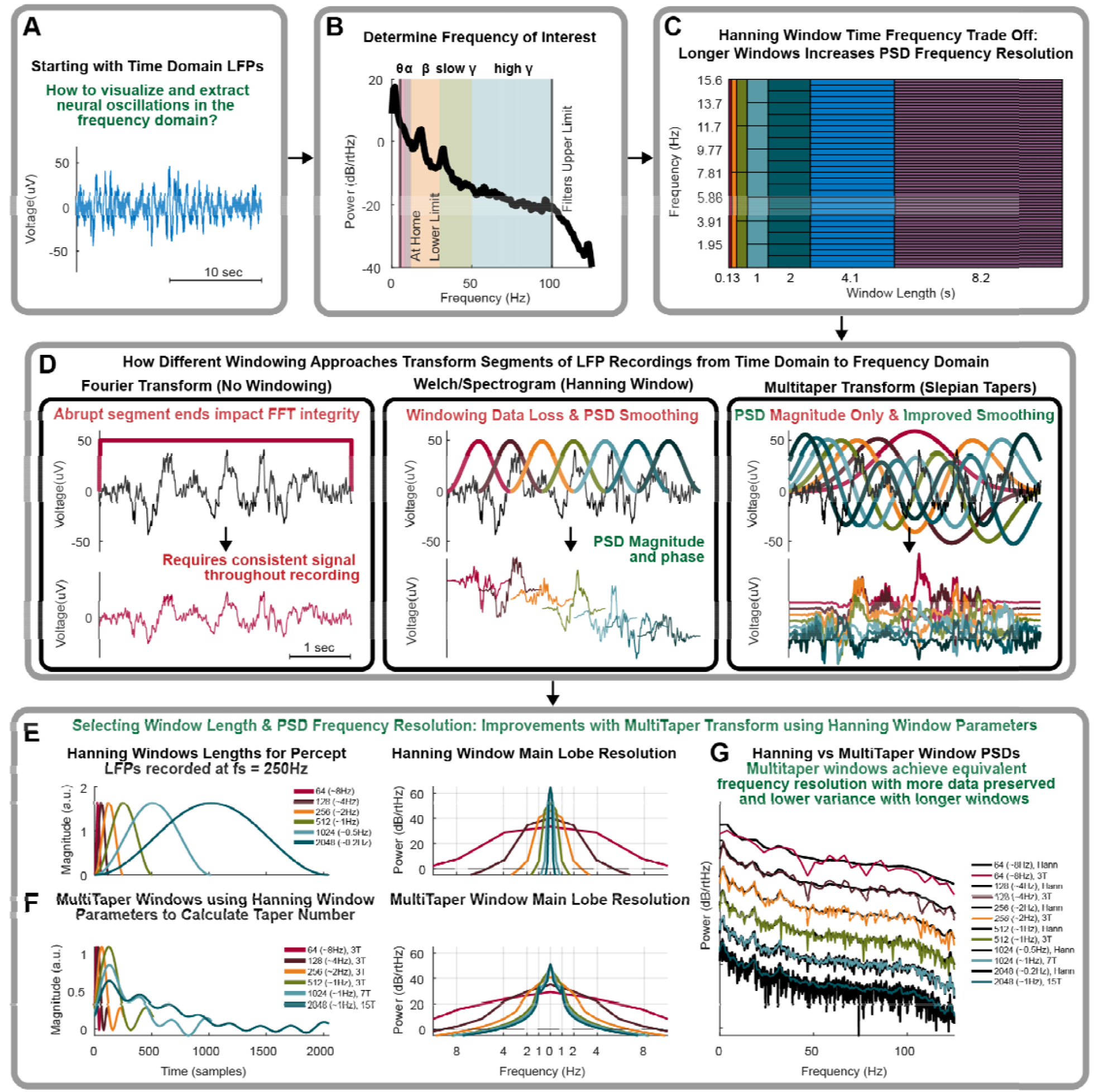
Frequency Transform Approaches of Neural Oscillations. **A) Time domain filtered LFP recording. B) Determine frequency bands of interest within data. Theta ϴ (4-8Hz), alpha ⍰(8-12Hz), beta β (12-30Hz), slow gamma γ (30-50Hz), or high gamma γ (50-100Hz). High gamma is limited by Percept™’s observable broadband. C) Hanning window Fourier transform tradeoff between the length of window, minimum frequency observable, and frequency precision due to PSD smoothing by the window. The x-axis shows time in samples and seconds. The shown blocks step in time increments of 32,64,128,256,512,1024, and 2048. D) Left: The Fourier transform with a full length rectangular window (no windowing). Middle: A segmented LFP recording with 50% overlapping Hanning windowing which can extract PSDs over time through a Spectrogram or be averaged overtime as with Welch’s method. Right: Multitaper transform approach using 7 Slepian tapers which are averaged together to construct a PSD. E) Hanning window lengths from 2^6 to 2^11, window lengths options for Percept™ LFPs sampled at 250Hz. The legend parenthesis frequency resolution is the main lobe half-bandwidth smoothing which is graphically shown in the right plot. F) Multitaper windows matched to the length and main lobe half-bandwidth smoothing resolution of the Hanning window for taper number determination. The legend and right figure are formatted similarly to Figure 2E. The longer length windows Multitaper smoothing resolution is held to 1Hz as the Hanning window resolution drops below 1Hz to increase taper count. G) The PSD of Hanning versus Multitaper windowing resolution and variance across different window lengths. Black is the Hanning window approach and the colored signal is the Multitaper approach for each window.**

First, the window length needs to capture the minimum frequency you plan to record. The sampling theorem indicates that the maximum frequency observed within a length of data requires two points from 1 period of the frequency, which is called the Nyquist frequency. Similarly, the minimum frequency observed requires 1 period of the frequency to occur within the window of data.

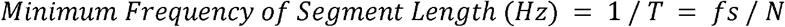

where T is the length of the window in seconds, *fs* is the sampling rate, and N is the length of the window in samples.

There is a trade off in resolution between frequency and time, as illustrated in Figure 2C. The Fourier transform frequency bin size which dictates the Discrete Fourier Transform (DFT) resolution between two frequencies is defined as:

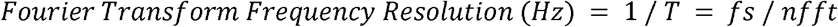

where `nfft` is the length of the Fourier transform. For example, using an `nfft`=256 samples and sampling rate of 250Hz, the minimum frequency is 1/T = *fs/*`nfft` ≈ 1Hz. The DFT then compares sine and cosine signals increasing in frequency of ∼1Hz. So, here the output frequency vector in a software implementation of the DFT would appear to have a resolution of ∼1Hz.

However, Figure 2C demonstrates the tradeoff shown due the frequency resolution smoothing induced by windowing the segment of data prior to computing the DFT. While a Fourier transform has a theoretical frequency resolution, the effective resolution is impacted by the window function and limits the resolution to:

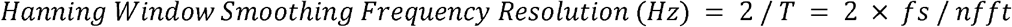

Using the example above, the DFT frequency resolution is ∼1 Hz. However, while the DFT may display 1Hz resolution, the Hanning window has a 2Hz smoothing effect, also known as the main lobe half-bandwidth, over the PSD. So, for example, this smoothes 10 Hz data between 8Hz and 12Hz, 11Hz data between 9 Hz and 13Hz, and so on. Understanding the impact of the window on the PSD is important when selecting a window length and estimating the frequency content of the LFPs.

### Multitaper Windowing

An alternative method for windowing LFP data is the multitaper method, which utilizes many orthogonal Slepian windowing functions. Multiple tapers can extract more information from the same window while maintaining the frequency resolution smoothing of the Hanning window. Figure 2D shows the Fourier transform (PSD), the Hanning window Welch method Fourier transform (PSD), the Hanning window over time Short Time Fourier Transform method (Spectrogram), and the Multitaper Transform (PSD). This illustrates the difference between not windowing, windowing using the Hanning method, and windowing using slepian tapers. As shown in the center plots of Figure 2D, smaller windows can be employed for improved temporal resolution at the cost of frequency resolution.

When generating a PSD, it is assumed that the frequencies of interest are present throughout the entire recording, which is often referred to as a stationary signal. Figure 2D illustrates how multiple slepian tapers (2D-right) utilize greater amounts of data when windowing, as compared to Hanning windows (2D-middle). While the combination of all Hanning windows may span the entire recording, one window is the limiting factor for frequency resolution. Whereas each Taper spans the entire signal, providing greater frequency resolution, and every additional taper allows for reduction in variance of the PSD through averaging.

Figure 2E shows multiple Hanning windows of varying lengths based around the sampling rate of Percept™ (250Hz). The main lobe width of these windows is shown by Fourier transforming each Hanning window. The main lobe width demonstrates how much smoothing each frequency affects adjacent frequencies. This smoothing is often quantified as the main lobe half-bandwidth, or half the main lobe width. For a Hanning window length of 256 samples, the main lobe width would be 4 Hz with a half-bandwidth of 2 Hz.

When conducting the Fourier transform of a Hanning windowed LFP segment, the frequency resolution or smoothing is defined by the window length. In a multitaper transform, the number of tapers is proportional to the smoothing or frequency resolution. Increasing the length of the signal allows for more tapers which lowers the PSD variance without sacrificing the frequency resolution of the transform. The number of tapers can be calculated as:

| Time half-bandwidth in terms of samples | Time half-bandwidth in terms of Hertz |
| --- | --- |
| $\text{Half Bandwidth (Hz)} = \Delta f \text{ (Hz)} / 2$ $W \text{ (cycles/sample)} = \text{Half Bandwidth (Hz)} / f_s \text{ (Hz)}$ $NW = N \text{ (samples)} \times W \text{ (cycles/sample)}$ | $\text{Half Bandwidth (Hz)} = \Delta f \text{ (Hz)} / 2$ $TW = T \text{ (seconds)} \times \text{Half Bandwidth (Hz)}$ |

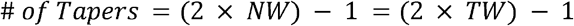

where *Δf* is the main lobe width in Hertz.

Multitaper windows are shown in Figure 2F and match the half-bandwidth and window length of Hanning windows in 2E with the corresponding number of tapers. As the length of the Hanning window increases, this allows for a greater number of tapers in the Multitaper window. In Figure 2F, the multitaper windows were Fourier transformed to demonstrate the main lobe width. These main lobe widths represent the maximal number of tapers while maintaining a 1Hz half-bandwidth at longer window lengths, which is why they appear wider than in Figure 2E. Using a longer Hanning window results in higher resolution and lower frequency smoothing. But, given the neural signals we are analyzing, as the Hanning window resolution drops below 1Hz, the Multitaper resolution can be maintained at 1Hz. This allows for acceptable frequency resolution but a further increase in the number of tapers which ultimately results in a cleaner PSD estimate.

Lastly, the PSDs determined using different Hanning (black) and Multitaper (colored) window lengths are shown in Figure 2G. For both approaches, longer window lengths results in less smoothed PSD frequency resolution. However, at the cost of phase data, Multitaper transforms extract more data through strategic windowing and result in cleaner PSDs by calibrating the frequency resolution to minimize the variance in the PSD.

### Artifact Rejection and Stationary Power Spectral Density Responses

When analyzing ANT LFPs, we aim to characterize background neural activity, which can be obscured by transient events such as movement artifacts or brief epileptiform episodes. PSDs are traditionally employed to assess persistent signal activity over the duration of a recording. However, large-amplitude transients can distort the entire PSD estimate, even when these events occur only in a short segment of the analyzed data. Here, we present several approaches to separate these transient events from the background activity within a recording. One approach to isolating background neural activity from transient events involves segmenting the LFP recording into smaller windows, removing outlying windows based on their spectral characteristics, and averaging the remaining segments. Alternatively, computing the principal components across these windows often reveals that the first principal component predominantly represents the background activity.

Typically, the segmentation of an LFP recording into smaller windows for frequency-domain analysis is performed using the short-time Fourier transform, which is represented as a spectrogram. Because phase information is not needed, a short-time Multitaper transform approach with 2 second windows, 50% overlapping windows, a half-bandwidth of 1Hz, and 3 total Slepian tapers per window. An example LFP recording is shown in Figure 3A with two large transients from stimulation and an epileptiform transient in the middle. The corresponding short time Multitaper spectrogram and individual PSD windows are displayed in Figures 3B and 3C.

**Figure 3:**
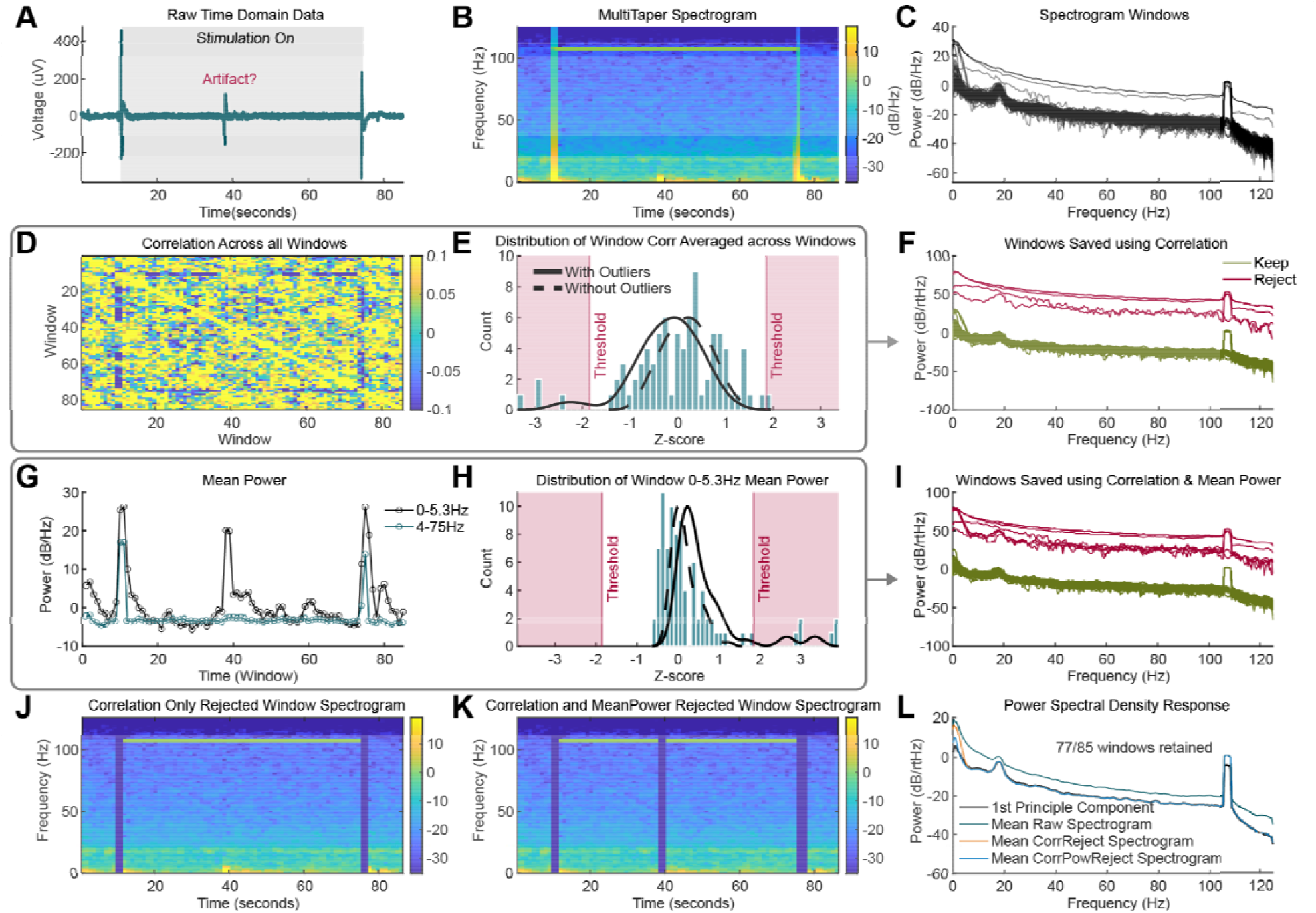
Nonstationary Signal Rejection Approaches. **A) Example time domain segment recorded using Percept where one stimulation setting was tested. There is roughly 10 seconds of baseline (stimulation off) activity before and after stimulation. Here we have a transient peak in the data that could be a form of interictal activity, an artifact, or some other pathological signal. B) The Multitaper spectrogram of the data is shown C) as well as all PSD windows plotted within the spectrogram. D) The correlation between all PSD windows is shown E) along with the distribution of the average correlation of one window with all other windows. That correlation is then z-scored and the outlier rejection threshold is shown based on the Grubbs test. A kernel density function shown with and without outliers representing the distribution of the data. F) Kept and rejected PSD windows based on their correlation alone are shown to demonstrate which signals will make up the mean PSD. G) The mean broadband (4-75Hz) and low frequency (0-5.3Hz) power for each window. H) The z-scores of the low frequency (0-5.3Hz) power for all the windows, with the outlier rejection shown in red and the kernel density function with and without outliers representing the distribution. I) The PSD kept and rejected windows are shown using the combined correlation and 0-5.3 Hz low frequency mean power rejection method. J,K) Based on rejected windows shown in F and I, the spectrogram in B is shown with the windows that were removed using J) correlation threshold rejection and K) the combination of correlation and low frequency mean power threshold rejection. L) When averaging across each spectrogram, the resulting PSD for the correlation only and correlation plus mean power filters are plotted against the raw PSD and first principal component of the signal PSD windows.**

First, we implemented a spectrogram window rejection filter by detecting outliers in the mean Pearson correlation of each broadband (4–75 Hz) PSD window with the remaining windows (Figure 3D,E). Using the Grubbs test, outliers were determined using an absolute value z-score of the correlation with a threshold set at ⍰=0.05. This approach corrects for variation in the number of spectrogram windows which is useful for datasets with varying total windows. Figure 3E shows a histogram and kernel density function of the z-score.

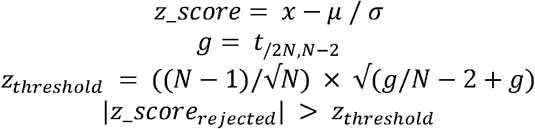

where N is the number of windows and arbitrary variable g represents t^2^_([⍰/2N),N-2_ which is the t-distribution threshold with N-2 degrees of freedom and ⍰/2N significance. t^2^_([⍰/2N),N-2_ can be calculated using the MATLAB `tinv` function.

This threshold approach identifies outliers by their correlation with other windows. The rejected windows from the individual PSD windows in Figure 3C are illustrated in Figure 3F. However, some windows containing transient signals may exhibit high-frequency broadband activity similar enough to other windows to avoid rejection by this correlation threshold, and thus certain artifact windows may persist.

PSD windows contaminated by movement or transient neural activity may exhibit large amplitude low-frequency components, making it beneficial to apply another rejection filter based on low-frequency power (0–5.3 Hz) rather than on broadband power (4–75 Hz), as shown in Figures 3G. To address these windows that might not be flagged by the correlation filter alone, an additional outlier rejection filter based on mean low-frequency power was implemented. We rejected windows with very low or high mean power (⍰=0.05), as illustrated in Figure 3H. The subsequent PSDs that were kept and rejected are shown in Figure 3I.

The individual PSD windows rejected based on the correlation threshold (Figure 3J) and the combined correlation and mean low-frequency power thresholds (Figure 3K) are shown. The background activity LFP PSD is then derived by averaging the PSD responses across the remaining time windows. By excluding windows that are dissimilar, or exhibit significantly different low-frequency power, the resulting PSD reflects an average of many similar windows with a consistent power spectral profile. While this example only rejects 8 windows, a few high deciBel windows can have a non-linear impact which will dominate the PSD response as shown in Figure 3L.

An alternative approach to the threshold rejection method involves analyzing the variance of each window in the spectrogram. Principal component analysis (PCA) can be used to identify spectral characteristics common to all windows. Since each window is normalized by its mean, the first principal component represents the most prominent variance observed. In LFP recordings where the majority of windows are not contaminated, this approach produces results similar to those obtained using the correlation and low-frequency mean power combination threshold rejection method, as shown in Figure 3L. However, if window similarity decreases across the recording, accurately extracting a PSD response with either method becomes more challenging.

Figure 3L illustrates the effect of each artifact removal approach on the final PSD. Removal of the stimulation and interictal artifacts from the LFP recording in Figure 3A—whether via principal component analysis or the combined correlation and mean power threshold rejection—enhances the visualization of the 20 Hz beta peak. It is evident that windows containing stimulus artifacts or transient activity—even if these artifacts are temporally localized—can affect the entire PSD estimated from those windows.

### Simplified 1/f Detrending Approach

Neural environmental pink noise—characterized by the 1/*f* dropoff in the PSD background—can complicate the estimation of oscillatory power across different frequencies. Here, we demonstrate one approach to decompose an LFP PSD into its aperiodic (1/*f*) and periodic components. It is important to note that this environmental pink noise is challenging to remove, and neural activity with amplitudes at or below this threshold remains indistinguishable, effectively serving as a local noise floor.

To detrend the 1/f aperiodic component, we implemented a method based on the logarithmic function defined in the Fitting Oscillations and One Over F (FOOOF) method [10], as shown in Equation 1. The raw time-domain LFP data and its multitaper power spectral density are presented in Figures 4A and 4B.

**Figure 4:**
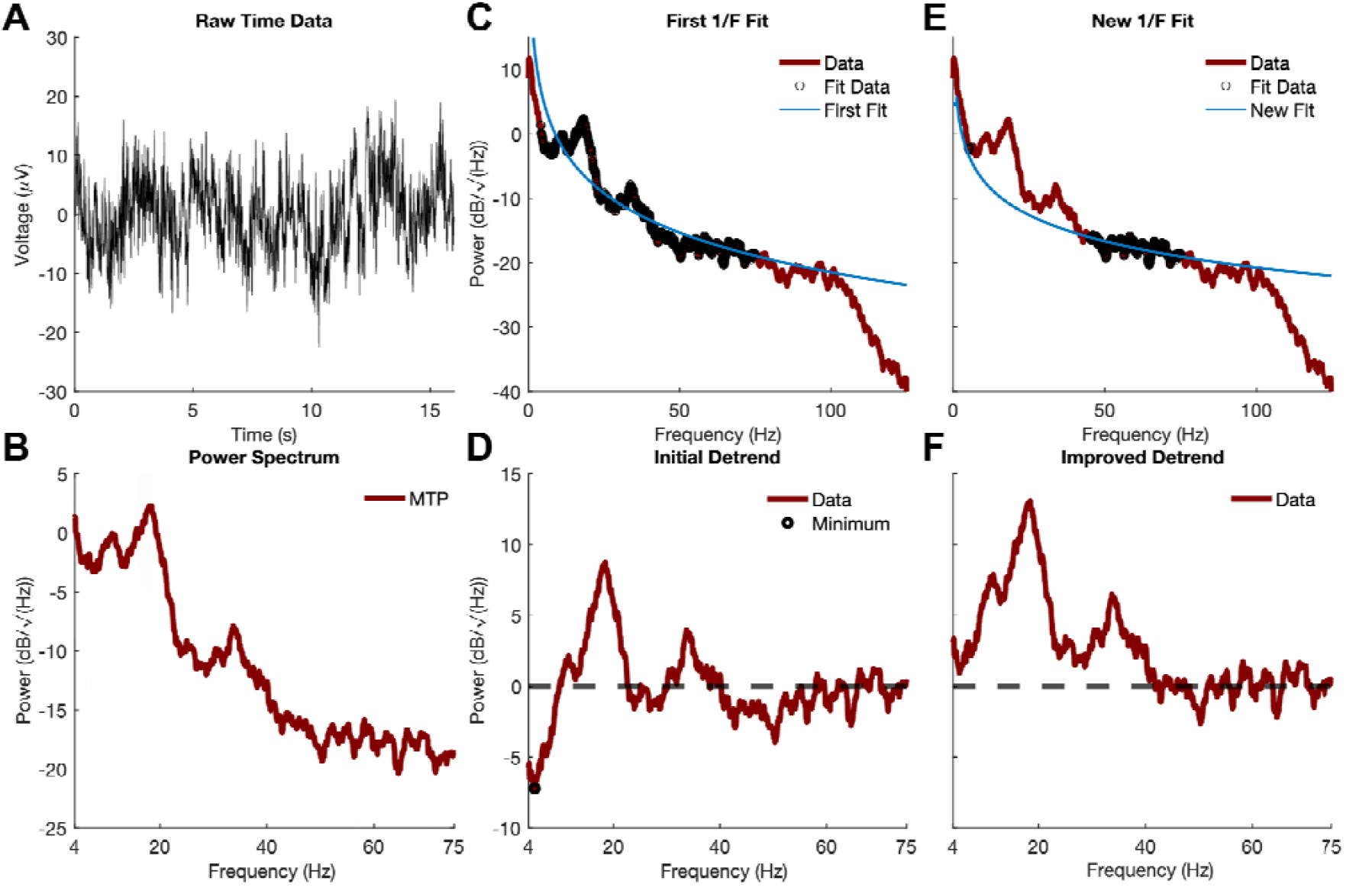
Detrending 1/*f* background neural activity. **A) Raw segment of time domain data recorded using Percept™. B) Frequency domain transform of time domain data using a Multitaper transform. C) An initial 1/*f* fit is extracted using the predefined broadband region of the data (4-75Hz). D) The detrended PSD after using the initial fit is used to identify minimums in the PSD which result from a fit of the total PSD not just 1/*f* aperiodic component. E) Using high broadband data points and the minimum points determined, a new fit is extracted from the data. F) Improved 1/*f* detrended signal to maintain as much area under the periodic frequency band components.**

where k is an x-axis shifting term, often referred to as the knee parameter, which defines the frequency at which the logarithmic function bend is observed in 1/*f*, b is the y-axis offset which shifts the entire function vertically, and x accounts for the decay of the logarithmic function.

To start, using a least squares regression fit, the function is fit to the PSD using the broadband power 4-75Hz, as shown in Figure 4C. This fit using the broadband data provides a good initial fit to the PSD. However, observed periodic activities (e.g., alpha, beta, gamma bands) often distort this fit by shifting it vertically. Consequently, the resulting detrended signal may contain both negative and positive values and a less prominent peak, as shown in Figure 4D. Since we assume the 1/f aperiodic component represents the noise floor—where activity below this level is not observable—we aim to preserve all activity above this floor. In an ideal scenario, the detrended signal would contain only positive values. To achieve this, we perform a second regression using a higher broadband frequency range, outside the periodic components, and a small frequency range determined by a tuned minimum search of the initial detrended PSD (Figure 4D). The new 1/f fit is less influenced by the periodic data, as demonstrated in Figure 4E, resulting in a detrended PSD that better preserves the neural periodic activity as shown in Figure 4F.

## Discussion

Despite device hardware and software filtering, noise can remain, and these noise sources may vary from patient to patient. Specifically, we have developed preprocessing approaches for Percept™-recorded LFPs to remove cardiac artifacts, transient non-stationary events, and aperiodic neural activity that can corrupt or confound the underlying neural signal.

In our trial, all participants implanted with Legacy leads exhibited cardiac artifacts in their LFP recordings at the first research visit. Because the Percept™ IPGs were retrofitted with Legacy leads that were not originally designed for neural recording, these artifacts were expected. In contrast, among participants implanted with Sensight™ leads, cardiac artifacts were observed in 50% of cases, with the first observable QRS complexes appearing around month 15 in three out of four Sensight™-implanted participants showing cardiac artifact.

There are multiple approaches to removing ECG artifacts. In many *in vivo* experimental setups, the ability to record the ECG in parallel with neural data provides a useful means for time-locked artifact filtering. However, the fully implanted nature of the Percept™ system precludes this straightforward approach, as any external system would require additional equipment and a mechanism for precise time locking to an external ECG trace. So, utilizing an approach to extract this signal from the LFPs when observed is preferable.

The presence of ECG artifacts may stem from several device-related factors. Historically, it is believed that fluid ingress at the implanted pulse generator’s bore connection of the lead extension had resulted in local cardiac signals being recorded alongside the neural signal in the subclavicular region at the implanted device [11–13]. Since Percept™ neural activity is captured via a differential amplifier that computes the difference between two recording electrodes [13], fluid ingress likely introduces an imbalance in artifacts on the electrodes, amplifying the observed cardiac artifact. Recent patents on the current Percept™ bore seal claim improvements that may mitigate this fluid ingress [11,13,14]. However, further investigation of the Percept™ system and leads over longer implant durations is necessary to fully characterize the causes of these cardiac artifacts.

It has also been shown that ECG artifacts can vary between stimulation on and off states, with LFP recordings exhibiting different levels of contamination depending on the sensing mode employed [15,16]. Other studies have examined the influence of implant region and chest placement of the neurostimulator on the likelihood of cardiac artifact occurrence in LFP recordings [17,18]. For instance, a 3.2-fold increase in ECG contamination was observed in left subclavicular implants (48.3%) compared to right-sided implants (15.3%) [17].

While understanding the root causes of cardiac artifact is important for long-term device design, our results indicate that LFP recordings can be recovered and analyzed using the cardiac artifact removal approach we demonstrated. Without removing the QRS complex, even small-amplitude cardiac artifacts can significantly contribute to the PSD due to their regularity, thereby confounding interpretation of neural activity, particularly in lower frequency bands.

Transient signals in the time-domain LFP data, such as movement artifacts or non-stationary neural activity, can manifest in various ways in the frequency domain. Thus, it is necessary either to remove these signals in the time domain or to reject the entire window in which the noise occurs in the frequency domain. Short-time frequency transform approaches that allow for the rejection of individual PSD windows are valuable, as they enable the isolation of consistent spectral features. In contrast, applying a Fourier transform to the entire recording makes it difficult to determine whether a particular frequency band reflects sustained activity or is dominated by a brief, high-amplitude transient event.

In the context of ANT DBS LFP recordings, the presence and role of interictal discharges (IEDs) within the ANT has been investigated [19,20]. These discharges are short-duration, high-amplitude events that primarily affect the lower frequency range of the PSD, while also generating harmonics at higher frequencies. Scalp EEG studies have shown that while the IED peak duration is often less than 50 milliseconds, the overall duration of an IED can exceed 100 milliseconds [21].

The time-domain neural activity observed in ANT DBS epilepsy participants can vary substantially, depending on the individual’s seizure network and the number of observable IEDs. This variability motivated our use of a systematic threshold approach to remove IEDs from LFP recordings when necessary. While other groups have focused on characterizing IEDs, our trial seeks to characterize the transient activity and stationary response to various stimulation parameters separately. Consequently, filtering is required to eliminate artifacts or interictal activity that might disproportionately influence estimates of the background PSD under either stimulation-on or stimulation-off conditions.

Finally, it has been suggested that the slope of the LFP aperiodic component—often described by the 1/f response—characterizes the excitation/inhibition balance in Parkinson’s disease [22] or during epileptiform activity [23]. Further studies aimed at disentangling the contributions of the recording environment, device noise, and neural activity to the 1/f component would be valuable. Such work would ensure that observed changes in the aperiodic response can be attributed solely to neural activity rather than to multiple confounding factors.

## Conclusions

While the ability to simultaneously stimulate and record is critical for investigating neural responses to stimulation, characterizing the neural activity from device-recorded LFPs requires significant preprocessing. Although Medtronic’s Percept™ system employs hardware and software filters to remove multiple noise sources, in-clinic LFPs collected via BrainSense may still contain cardiac artifacts. Furthermore, when analyzing LFPs in the frequency domain, the removal of transient neural activity and aperiodic components enhances the accuracy of neural oscillation comparisons. Preprocessing is not a one-size-fits-all solution; it requires careful examination of each participant’s data to determine the necessity and sufficiency of noise and artifact removal.

## Acknowledgements

A special thank you to Scott Stanslaski for technical assistance when using the Medtronic Percept™ system within our ongoing clinical trial.

## Data Availability Statement

Data can be made available upon reasonable request to the corresponding author.

